# Major outbreak of endophthalmitis after cataract surgery: a retrospective cohort in northern Brazil

**DOI:** 10.1101/2023.02.08.23285557

**Authors:** Priscilla Perez da Silva Pereira, Andriely Alayne Carvalho Sabini, Rosa Maria Ferreira de Almeida, Daniela Oliveira Pontes, Márcia Maria Bezerra Mororó Alves, Viviane Alves de Sousa, Magzan da Silva Azevedo, Adalgiza de Souza Botelho, Surlange Freire Ramalhaes, Edilson Batista da Silva

**Author notes:** Endereço atual: ^#uma^Departamento de Enfermagem, Universidade Federal de Rondônia, Campus Ribeiro Filho, km 9,5, sentido Acre, CEP 76801-059. Porto Velho, Rondônia, RO, Brasil. **Corresponding Author** (AACS). **Author contribution**. Conceptualization: PPSP, AACS, RMFA, DOP. Data Curation: PPSP, AACS, RMFA, DOP, MMBMA, VAS, MSA, ASB, SFR, EBS. Formal Analysis: PPSP, AACS, RMFA, DOP. Investigation: RMFA, DOP, MMBMA, VAS, MSA, ASB, SFR, EBS. Methodology: PPSP, AACS, RMFA, DOP Project Administration: PPSP. Supervision: PPSP, AACS, RMFA, DOP, MMBMA. Writing – Original Draft Preparation: PPSP, AACS, RMFA, DOP, MMBMA, VAS, MSA, ASB, SFR, EBS. Writing – Review & Editing: PPSP, AACS, RMFA, DOP, MMBMA, VAS, MSA, ASB, SFR, EBS.

## Abstract

**Background:** Endophthalmitis is one of the most important adverse events after cataract surgery as it can lead to total vision loss. The aim of this study was to describe the occurrence of endophthalmitis after phacoemulsification with intraocular lens implantation among patients assisted during a joint effort in Porto Velho, Rondônia, Brazil.

**Method:** This is a retrospective cohort study, carried out from a bank with 649 medical records of patients who underwent surgery. Descriptive analysis and multiple analysis using Robust Poisson Regression were performed to estimate relative risks (RR) and 95% confidence intervals (95%CI). A statistical analysis was performed using the statistical program Stata® version 16.0 (College Station, Texas, USA).

**Results:** The incidence of postoperative endophthalmitis confirmed by culture was 10.88%, the highest ever recorded in the world. A higher risk for endophthalmitis was found, in probable cases, among males (RR: 1.88; 95%CI:1.03; 3.44) and brown and yellow skin color (RR: 2.78; 95 %CI %: 1.17; 6.60). For confirmed and probable cases, bilateral surgery and specific lens model were also risk factors. The predominant etiological agents were gram-negative and the main clinical manifestation was corneal edema. The average number of days to start treatment was eight days and 27.12% used antibiotics.

**Conclusion:** Specific protocols are needed for cataract surgeries that encompass hiring, performing and monitoring these services to ensure good practices and patient safety.

## Introduction

Cataracts are responsible for up to 50% of cases of blindness in the world^[1]^. It is defined as any clouding of the lens with or without a decrease in visual acuity. Advanced age, exposure to sunlight, comorbidities, previous eye diseases, ocular trauma, socioeconomic status and the region where the individual resides are factors that directly or indirectly interfere with the appearance of cataracts^[2]^.

To recover the visual capacity of the senile cataract carrier, surgery is the only option, since the treatment with prescription glasses is transitory. Facectomy, associated with the implantation of an intraocular lens, is a safe and effective procedure, providing visual rehabilitation in the vast majority of cases^[3]^.

One of the main complications of facectomy is endophthalmitis. Its clinical presentation usually occurs between one and seven days after surgery, but following the recommendations of the Brazilian Ministry of Health, surveillance in search of symptoms and symptoms suggestive of infection must occur within 90 days after the procedure when lens implants were used^[4]^. The main signs and symptoms of endophthalmitis are conjunctival hyperemia, hypopyon, cloudy vitreous, anterior chamber reaction, corneal edema and loss of visual acuity^[4]^.

Endophthalmitis must be recognized early, as in severe cases it can lead to blindness^[5]^. In addition to clinical consequences, there are also consequences in relation to the individual’s functional capacity, such as difficulty in carrying out daily activities and mental outcomes such as isolation, sadness, irritability and even depression^[3]^. As for health service costs, a study conducted in the United States of America found that the occurrence of endophthalmitis after cataract surgery increased 83% of expenses with reimbursements when compared to those who did not develop the complication, which corresponded to the year from 2014 to $4,893 USD^[6]^.

In most cases, endophthalmitis is caused by the presence of bacteria found on the palpebral margin and tear film of the patient’s conjunctival flora^[7]^. Other possible sources of infection may be the contamination of equipment, instruments, supplies, solutions, surgical drapes, lenses, or lack of good practice by health professionals^[7, 8]^.

According to a study carried out in Poland, after more than one million facectomy surgeries, in a period of five years, the incidence rate of endophthalmitis was 0.066%^[9]^. Another study conducted in Korea between 2014 and 2017, involving nearly one million patients, found an incidence of 0.063%^[10]^. In the United States, a survey of more than eight million cataract removal surgeries found an incidence of 0.04%^[11]^. In Malaysia, a cohort from 2008 to 2014 reported an incidence of 0.08%^[12]^. In Brazil, in a study conducted between 2008 and 2014, after more than 27,000 cataract surgeries, the incidence was 0.13%, with an annual variation of 0.04% to 0.27%^[2]^. Another Brazilian study conducted in a northern Brazilian state found that after 3,999 procedures performed in 2013 and 2014 in joint efforts, 1.67% had perioperative or postoperative complications^[13]^.

To compensate for the appearance of new cases of cataracts and reduce the alarming number of existing cases, it is estimated that it would be necessary to perform approximately 550,000 cataract surgeries in Brazil per year. In order to improve the access of the Brazilian population, mainly in the most distant regions, to minimize budgetary and managerial limitations, joint efforts are carried out^[13]^. Mutirão is the Portuguese term that refers to the organization of national campaigns for elective surgeries with the purpose of reducing queues and waiting time for procedures^[1]^.

In the period from 02/14/2022 to 02/23/2022 in the municipality of Porto Velho, one of the capitals of the northern region of Brazil, there was a joint effort of cataract surgeries. On 02/23/2022, in the outpatient care of patients undergoing procedures, the first suspected case of endophthalmitis was identified. On the same day, the surgeries were suspended by the company responsible for the task force. After initial investigation of the cases by the Center for Strategic Information on Health Surveillance of the State of Rondônia, an outbreak of endophthalmitis was confirmed. Thus, the aim of this study is to describe the occurrence of endophthalmitis after phacoemulsification with intraocular lens implantation among patients assisted during a joint effort in Porto Velho, Rondônia, Brazil.

## Method

Retrospective cohort study, carried out from a database with 649 medical records of patients who underwent phacoemulsification surgery with intraocular lens implantation in the city of Porto Velho, Rondônia, Northern Brazil. Some patients underwent bilateral surgery, so there were 1,044 procedures in a period of 10 days, with an average of 104 procedures per day (minimum 90 and maximum 120 procedures).

The State of Rondônia is located in the North Region of Brazil and is composed of 52 municipalities with an estimated population in 2021 of 1,815,278 inhabitants. About 73% of the state’s population lives in urban areas, the Human Development Index (HDI) was 0.690 and 6.02% of the population is over 60 years old^[14]^.

Information was collected from the medical records of users who underwent the Phacoemulsification procedure with implantation of an intraocular lens from February 14 to 23, 2022 during an ophthalmological surgeries campaign in Porto Velho. The information was typed into a standardized electronic form and the researchers were previously trained. The information collected was: sociodemographic; previous exams; past pathological history; materials and methods used in the procedure; signs and symptoms after the procedure; examinations and treatment after the procedure. Not all the collected variables were used in this initial analysis, it was decided to present in this article the variables with greater statistical or epidemiological relevance already known.

A probable case was defined as an individual undergoing the procedure who presented at least two of the following signs and symptoms: low visual acuity, ocular pain, corneal edema, conjunctival hyperemia, hypopyon, anterior chamber reaction, cloudy vitreous and/or patient submitted to postoperative intravitreal antimicrobial injection^[4]^. Individuals submitted to the procedure and with a laboratory diagnosis with a positive culture result in vitreous and/or aqueous humor were considered as confirmed cases^[4]^. The two case definitions were chosen because using only culture-proven cases could lead to underestimation of the reported true incidence^[15]^.

Data were presented in the form of absolute and relative frequencies and measures of central tendency were used for numeric variables. Pearson’s chi-square test (x^2^) and Fisher’s exact test were performed in the bivariate analysis. After bivariate analysis, the covariates were tested for the presence of multicollinearity, represented by correlations greater than 0.80. All variables with a significance level of 10% or epidemiological relevance were considered as adjustment variables and submitted to multiple analysis.

The multiple analysis performed was Poisson Robust Regression to estimate relative risks (RR) and 95% confidence intervals (95%CI) using the forward selection strategy. In the saturated model, variables were maintained with results of p<0.05 or that adjusted the association measure by at least 10%, relevant to the outcome in question or that improved the quality of the final model. Statistical analysis was performed using the statistical program Stata® version 16.0 (College Station, Texas, USA).

This study is part of the matrix project entitled: Good Practices in Patient Care, Infection Control and Processing of Health Products in the State of Rondônia, authorized by the Ethics and Research Committee of the Federal University of Rondônia, CAE 20070719.5.0000.5300.

## Results

There were 649 medical records analyzed, the incidence of confirmed endophthalmitis was 11.94% (95% CI: 9.43;14.85) and probable endophthalmitis 10.88% (95% CI: 8.46; 13.70).

Half of the patients were female; most declared their skin color as brown or yellow and were over 60 years old (Table 1). Some patients already had a history of previous surgery (22.72%), used antibiotics (19.34%), more than half underwent bilateral surgery, approximately half of the users received the Eyeol® model intraocular lens.

**Table 1.**
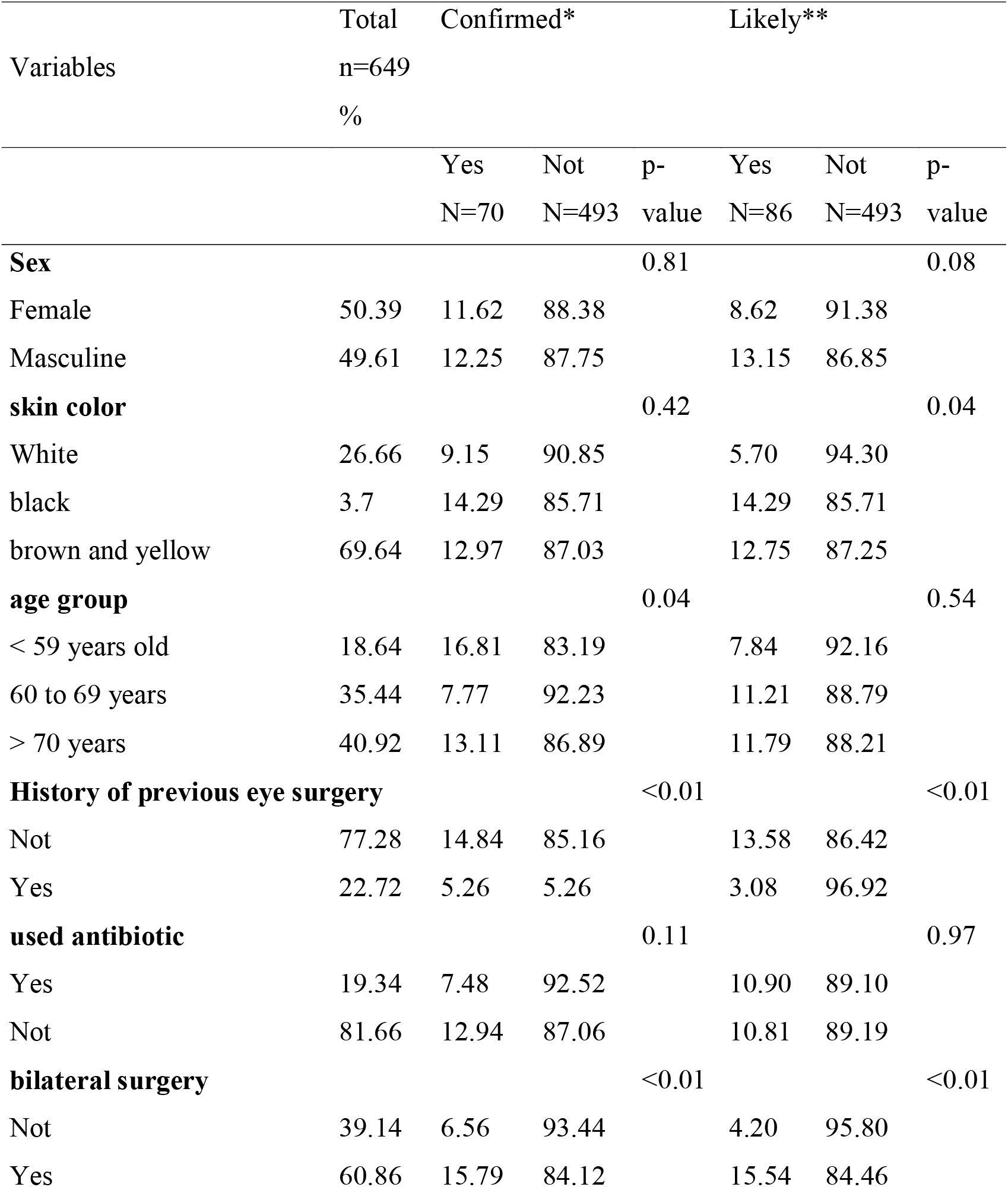
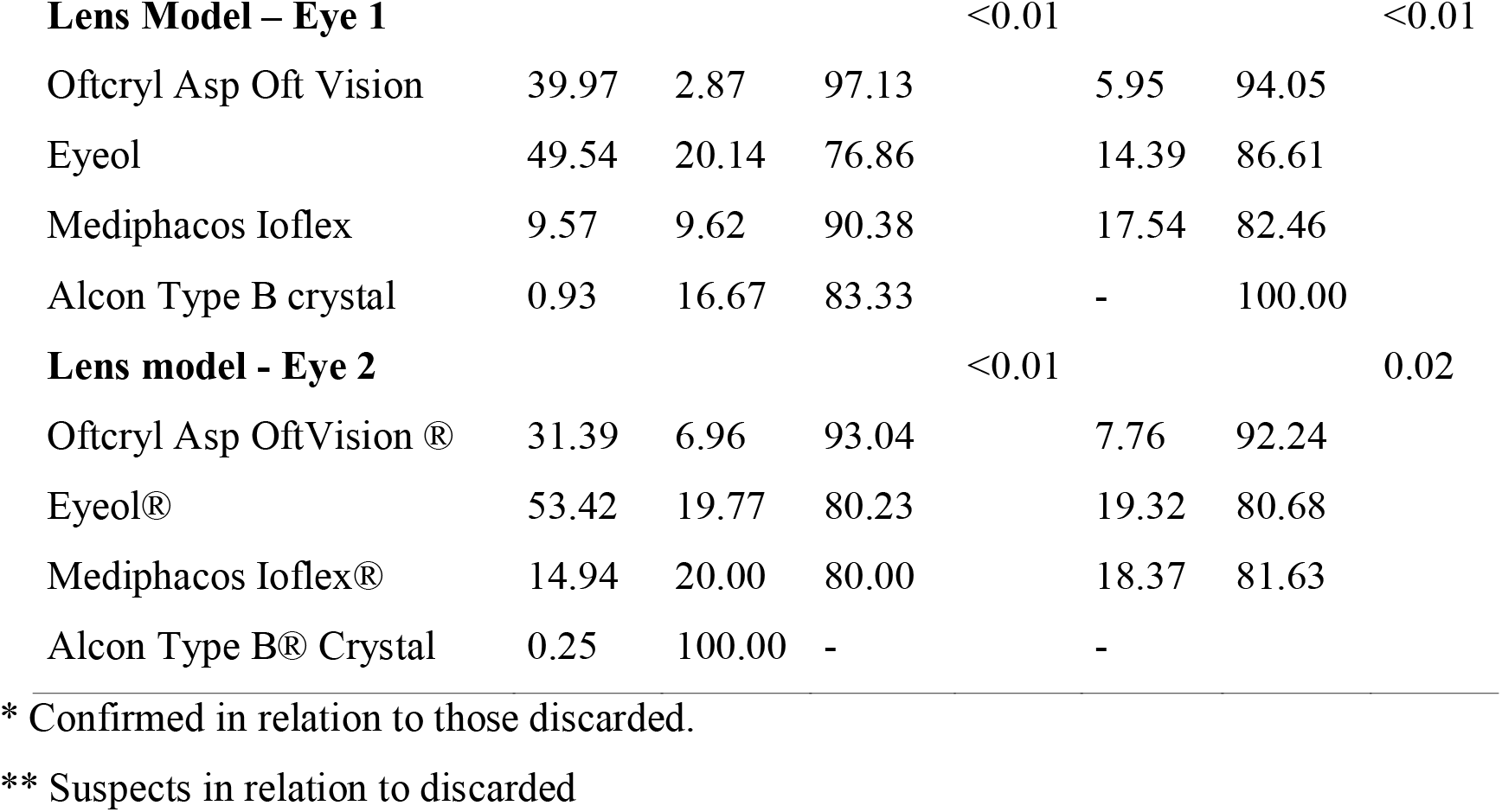
Characteristics of users undergoing Phacoemulsification with intraocular lens implantation, Porto Velho-RO, 2022 (n=649)

Statistically significant differences were found for confirmed cases in relation to age group, history of previous ophthalmological surgery, bilateral surgery and lens model. Regarding probable cases, skin color, history of previous ophthalmic surgery, bilateral surgery, lens model and completion of the safe surgery checklist were statistically significant.

For confirmed cases, in the crude analysis, age between 60 and 69 years was a protective factor, as well as having a history of previous ophthalmologic surgery (Table 2). Having surgery in both eyes increased the chances of endophthalmitis 2.4 times and some lens models had a higher risk for endophthalmitis. In the adjusted analysis, bilateral surgery presented collinearity with the variable of confirmed cases, the use of the Eyeol lens remained as a risk factor in eye 1 and the Cristal Alcon Type B lens presented itself as a risk in use in eye 2.

**Table 2.**
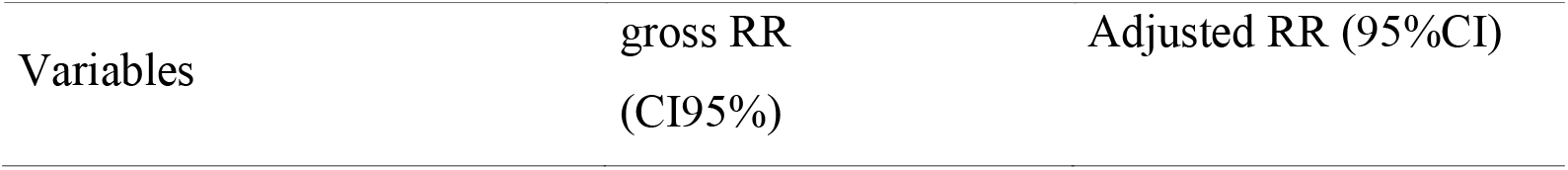
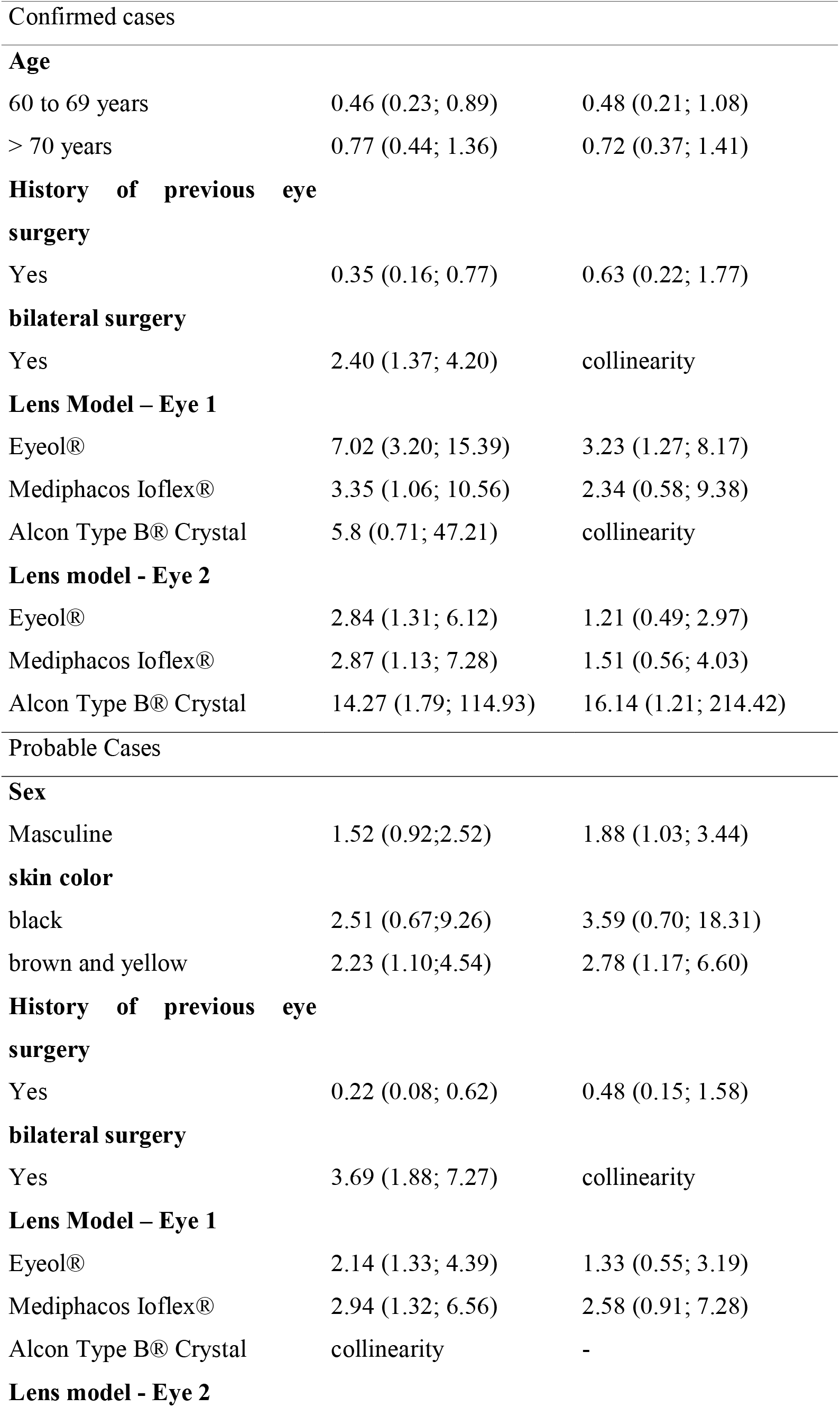
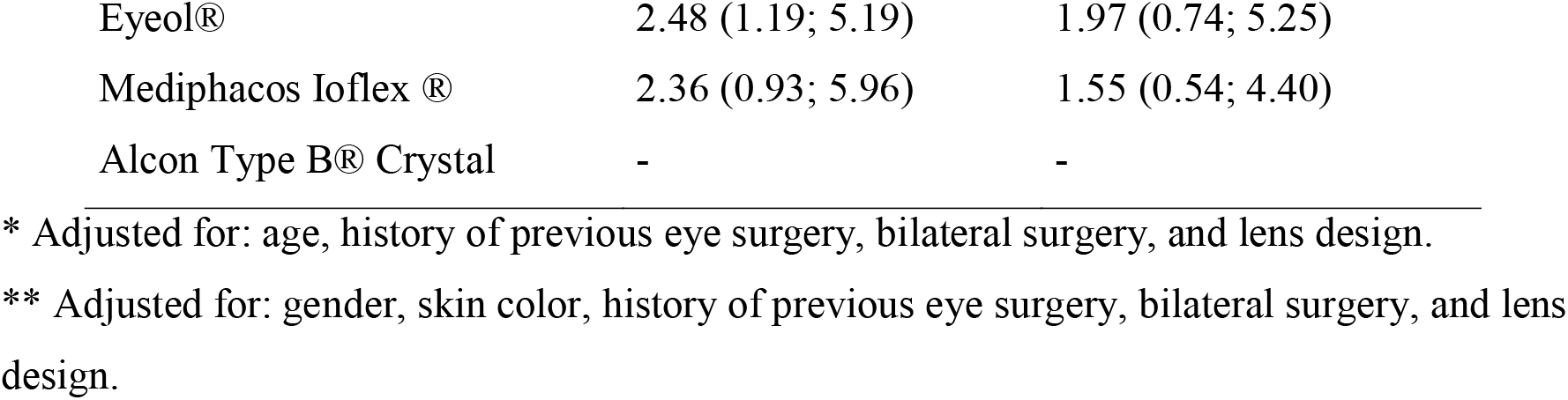
Relative risk analysis of the characteristics of users undergoing Phacoemulsification with intraocular lens implantation, Porto Velho, Rondônia, Brazil, 2022 (n=649)

| Variables | gross RR<br>(CI95%) | Adjusted RR (95%CI) |
| --- | --- | --- |
| **Age** |  |  |
| 60 to 69 years | 0.46 (0.23; 0.89) | 0.48 (0.21; 1.08) |
| > 70 years | 0.77 (0.44; 1.36) | 0.72 (0.37; 1.41) |
| **History of previous eye surgery** |  |  |
| Yes | 0.35 (0.16; 0.77) | 0.63 (0.22; 1.77) |
| **bilateral surgery** |  |  |
| Yes | 2.40 (1.37; 4.20) | collinearity |
| **Lens Model – Eye 1** |  |  |
| Eyeol® | 7.02 (3.20; 15.39) | 3.23 (1.27; 8.17) |
| Mediphacos Ioflex® | 3.35 (1.06; 10.56) | 2.34 (0.58; 9.38) |
| Alcon Type B® Crystal | 5.8 (0.71; 47.21) | collinearity |
| **Lens model - Eye 2** |  |  |
| Eyeol® | 2.84 (1.31; 6.12) | 1.21 (0.49; 2.97) |
| Mediphacos Ioflex® | 2.87 (1.13; 7.28) | 1.51 (0.56; 4.03) |
| Alcon Type B® Crystal | 14.27 (1.79; 114.93) | 16.14 (1.21; 214.42) |
| **Sex** |  |  |
| Masculine | 1.52 (0.92; 2.52) | 1.88 (1.03; 3.44) |
| **skin color** |  |  |
| black | 2.51 (0.67; 9.26) | 3.59 (0.70; 18.31) |
| brown and yellow | 2.23 (1.10; 4.54) | 2.78 (1.17; 6.60) |
| **History of previous eye surgery** |  |  |
| Yes | 0.22 (0.08; 0.62) | 0.48 (0.15; 1.58) |
| **bilateral surgery** |  |  |
| Yes | 3.69 (1.88; 7.27) | collinearity |
| **Lens Model – Eye 1** |  |  |
| Eyeol® | 2.14 (1.33; 4.39) | 1.33 (0.55; 3.19) |
| Mediphacos Ioflex® | 2.94 (1.32; 6.56) | 2.58 (0.91; 7.28) |
| Alcon Type B® Crystal | collinearity | - |
| **Lens model - Eye 2** |  |  |
| Eyeol® | 2.48 (1.19; 5.19) | 1.97 (0.74; 5.25) |
| Mediphacos Ioflex ® | 2.36 (0.93; 5.96) | 1.55 (0.54; 4.40) |
| Alcon Type B® Crystal | - | - |

\* Adjusted for: age, history of previous eye surgery, bilateral surgery, and lens design.
\*\* Adjusted for: gender, skin color, history of previous eye surgery, bilateral surgery, and lens design.

In relation to probable cases, in the crude analysis brown and yellow skin color was presented as a risk factor. A history of ophthalmological surgery as a protective factor and the use of Eyeol lenses in both eyes, Mediphacos Ioflex in eye 1 and Cristal Alcon Type B were a risk factor for ophthalmitis. In the adjusted analysis, brown or yellow skin color remained statistically significant and presented a risk 2.78 times (CI 95%: 1.17; 6.60) to develop endophthalmitis, previous history of ophthalmic surgery showed collinearity and none lens model showed significance

The average number of days between the date of the procedure and the date of initiation of treatment was eight days (minimum of 1 and maximum of 38 days). Of the total number of medical records evaluated, 97.69% had a consultation record up to 24 hours after the surgical procedure, and of these, 73.50% had some sign or symptom (Table 3). In the second consultation, 37.44% had symptoms, but in this second consultation, half of the patients returned for evaluation of the procedure performed on the second eye. In the third consultation 32.36% reported signs or symptoms, in the fourth consultation 19.88% and in the fifth consultation 13.56%.

**Table 3.** Frequency of signs and symptoms presented by time of return visit, Porto Velho, Rondônia, Brazil, 2022 (n=649)

| Signals and symptoms * | first<br>consultat | second<br>consultatio | Third<br>consultat | Fourth<br>appointm | Fifth<br>Query |
| --- | --- | --- | --- | --- | --- |

|  | ion<br>(n=634) | n<br>(n=392) | ion<br>(n=485) | ent<br>(n=185) | (n=99) |
| --- | --- | --- | --- | --- | --- |
| corneal edema | 68.26 | 31.12 | 12.33 | 14.79 | 9.09 |
| hypopyon | 1.08 | 1.69 | 12.94 | 3.39 | 2.16 |
| low visual acuity | 1.08 | 1.69 | 13.71 | 3.39 | 1.39 |
| conjunctival hyperemia | 0.31 | 0.31 | 3.85 | 10.94 | 6.93 |
| cloudy vitreous | - | - | - | 0.15 | - |
| eye pain | 0.31 | 0.15 | 0.31 | 1.39 | 0.62 |
| anterior chamber reaction | 5.24 | 2.62 | 10.32 | 10.02 | 5.39 |
| Others | 3.39 | 2.47 | 15.72 | 10.94 | 6.47 |

\* Patients may have presented more than one sign and symptom.
\*\* Other – the main symptoms recorded were inflammatory membranes, bullous keratopathy and hyposphagma.

Of the total number of medical records evaluated, 21.91% collected biological material. Three microorganisms were identified - *Pseudomonas luteola, Pseudomonas oryzihabitans and Acinobacter baumanni* in the collections of vitreous and/or aqueous humor (Table 4).

**Table 4.** Microorganism identified according to culture of biological material, Porto Velho, Rondônia, Brazil, 2022

| microorganism | Vitreous<br>humor | aqueous humor |
| --- | --- | --- |
| *Pseudomonas luteola* | 44 | 5 |
| *Pseudomonas oryzihabitans* | two | 1 |
| *Acinobacter baumannii* | 22 | 0 |
| there was no growth | 25 | 8 |
| Result not found | 42 | 61 |
| Total* | 135 | 75 |

\*Some patients performed more than one collection

According to the records, of the total of 649 medical records, 12.17% indicated the use of atropine, 14.79% prednisone, 17.57% vancomycin, 3.54% dexamethasone, 14.95% amikacin, in addition to of other medications with less than 2% of registrations.

## Discussion

Contrary to the world scenario, where the incidence of endophthalmitis is declining, the incidence of postoperative endophthalmitis in Rondônia was 10.88%, a result much higher than the incidence found in the literature, thus characterizing the largest outbreak of endophthalmitis after cataract surgery ever registered in the world^[12,16,9]^.

Regarding demographic characteristics, a higher risk for endophthalmitis was found, in probable cases, among males. This fact may be due to differences in the ocular flora or variable adherence to postoperative care, which can increase complications in cataract surgeries^[12,17,9]^.

When analyzing the skin color of the patients, the predominance of brown and yellow presented itself as a risk factor of 2.78 times (CI 95%: 1.17; 6.60) for probable cases of endophthalmitis when compared to people white skin. A study conducted in the United States of America between 2010 and 2014 with more than 150,000 patients found higher rates of endophthalmitis after cataract surgery among men, older individuals, African Americans and Native Americans^[6]^.

Having surgery performed on both eyes increased the odds of endophthalmitis 2.4 times among probable cases. A systematic review conducted with 14 studies involving 276,260 people concluded that there is a lack of evidence on the cost-effectiveness of performing ophthalmic procedures on the same patient on different days^[18]^.

The most common etiological agents identified in endophthalmitis after cataract surgery are gram-positive, especially negative *coagulase* Staphylococcus, followed by gram-negative and, less commonly, fungi^[19,20,21]^. A systematic review of studies conducted in India found that the most common microorganisms in endophthalmitis after cataract surgery were Staphylococcus, followed by Gram-negative bacilli of Pseudomonas species^[15]^. In our study, there was a predominance of gram-negative being among the main ones: *Pseudomonas luteola, Pseudomonas oryzihabitans and Acinobacter baumanni* in the collections of vitreous and/or aqueous humor. This fact is also found in two studies conducted in Asia^[5,22]^. However, endophthalmitis caused by other species such as Pseudomonas oryzihabitans, Pseudomonas stutzeri, and P. luteola are rare, and the first case of endophthalmitis caused by P. luteola was reported in Asia in 2018^[23]^.

The occurrence of contamination by microorganisms can be associated with different causes, such as, for example, contamination of the ocular viscoelastic device^[19]^, vials used in surgeries^[24]^, pre-filled syringes of saline solution^[25]^. Or it may be related to the surgeon’s experience^[6]^, number of surgeries performed^[6]^ and failures in work processes, with emphasis on the inadequate processing of surgical instruments^[4]^.

The main clinical manifestation of endophthalmitis was corneal edema with a proportion of 68.26% patients, in the first consultation after the surgical procedure, followed by hypopyon, low acuity, conjunctival hyperemia, ocular pain, anterior chamber reaction and other symptoms. Eye pain and hypopyon occur in up to 75% of clinical manifestations of endophthalmitis after cataract surgery^[5,22]^.

The average number of days between the date of the procedure and the date of treatment initiation was eight days, a result similar to that reported in the literature^[5,26,27]^. In addition, within this period, there must be availability for the patient’s prompt care at any time, in case of need^[5]^. Most complications after cataract surgery occur in the late postoperative period, which reinforces the importance of actively seeking out patients after the procedure, even in cases of joint efforts - which does not always occur in this type of care in Brazil^[13]^.

The conduct of using atropine, prednisone, vancomycin, dexamethasone and amikacin is in accordance with the literature, as it is recommended that the treatment of endophthalmitis be based on the selection of effective and safe antimicrobials. A broad-spectrum therapy for gram-positive and gram-negative organisms, such as vancomycin and intravitreal ceftazidine or amikacin, is initially suggested.

In order to avoid the adverse event, there are some registered preventive actions for endophthalmitis, such as the use of 0.66% povidone-iodine eye drops before surgery^[28]^. Or use of intracameral antibiotics during surgical procedures^[29,21]^ can be considered as a preventive approach to infections.

Among the limitations of this study is the fact that it was not possible to verify potential sources of infection such as instruments, lenses, surfaces or others. The data used in this study was taken from the medical records filled out by the professionals hired for the task force, which may have flaws in filling in information and defining cases due to the absence of requests for microorganism culture tests, for example. Another limitation was the fact that it was not possible to verify how postoperative care was given by the patients themselves in their homes.

As this is the largest outbreak of endophthalmitis ever recorded in the world, it is understood that the results are useful to draw attention mainly to the need for specific protocols for contracting, carrying out and monitoring care services of the joint effort type of cataract surgeries, common practice mainly in regions with little access to health services.

During the outbreak investigation, all professionals involved in the task force were interviewed by the state health surveillance team. These interviews were transcribed and will be analyzed to find flaws in the process of conducting surgeries in all its stages - pre, peri and post aiming at creating mechanisms that can regulate the occurrence of joint efforts for cataract surgeries, as well as promoting the training of health care professionals involved directly with patient care or indirectly as in the preparation of instruments used in the procedure and health surveillance professionals responsible for monitoring the actions carried out by health institutions.

## Supporting information

Acknowledgment

## Data Availability

All relevant data are within the manuscript and its Supporting Information files.

