## Supplementary material for "Major outbreak of endophthalmitis after cataract surgery: a retrospective cohort in northern Brazil": Acknowledgment

To Agevisa employees, who contributed to data collection and investigation of the outbreak....

- Gilmar Meireles Nogueira - Specialist Nurse Infection Control Coordination

- Luma Akemi de Azevedo Kubota - Center for Noncommunicable Diseases and Injuries

- Rosiane Maciel Batista Ximenes – Non-Biological Risks Center

- Alessandra da Silva Dantas - polio coordinator

- Ana Lúcia Teles – NSS/GTVISA

- Letícia Aline Ricci - State Coordinator of Death with defined underlying cause

- Liziane Sandra Silva Mendonça - State Coordinator of Child and Fetal Death Surveillance

- Cesarino Júnior Lima Aprígio - Environmental Surveillance Technical Manager

- Maria Leiliane de Brito - Technical Manager of Health Surveillance

- Maria Arlete da Gama Baldez - Technical Manager of Epidemiological Surveillance
